# Quantification of seizure termination patterns reveals limited pathways to seizure end

**DOI:** 10.1101/2021.03.03.21252789

**Authors:** Pariya Salami, Mia Borzello, Mark A. Kramer, M. Brandon Westover, Sydney S. Cash

## Abstract

Seizures result from a variety of pathologies and exhibit great diversity in their dynamics. Although many studies have examined the dynamics of seizure initiation, few have investigated the mechanisms leading to seizure termination. We examined intracranial recordings from patients with intractable focal epilepsy to differentiate seizure termination patterns and investigate whether these termination patterns are indicative of different underlying mechanisms.

Seizures (*n*=710) were recorded intracranially from 104 patients and visually classified as focal or secondarily generalized. Only two patterns emerged from this analysis: (a) those that end simultaneously across the brain (synchronous termination), and (b) those whose ictal activity terminates in some regions but continues in others (asynchronous termination). Finally, seizures ended with either an intermittent bursting pattern (burst suppression pattern), or continuous activity (continuous bursting). These findings allowed for a classification and quantification of the burst suppression ratio, absolute energy and network connectivity of all seizures and comparison across different seizure termination patterns.

We found that different termination patterns can manifest within a single patient, even in seizures originating from the same onset locations. Most seizures terminate with patterns of burst suppression regardless of generalization but that seizure that secondarily generalize show burst suppression patterns in 90% of cases, while only 60% of focal seizures exhibit burst suppression. Interestingly, we found similar absolute energy and burst suppression ratios in seizures with synchronous and asynchronous termination, while seizures with continuous bursting were found to be different from seizures with burst suppression, showing lower energy during seizure and lower burst suppression ratio at the start and end of seizure. Finally, network density was observed to increase with seizure progression, with significantly lower densities in seizures with continuous bursting compared to seizures with burst suppression.

Our study demonstrates that there are a limited number of seizure termination patterns, suggesting that, unlike seizure initiation, the number of mechanisms underlying seizure termination is constrained. The study of termination patterns may provide useful clues about how these seizures may be managed, which in turn may lead to more targeted modes of therapy for seizure control.

## 1 Introduction

Seizures can stem from a multitude of etiologies and manifest a wide variety of electrographic dynamics.^1–3^ Over the past few decades, different aspects of the electrographic manifestation of seizures have been studied with the goal of understanding mechanisms underlying seizures. Most of these studies have focused on seizure initiation; seizure termination has been correspondingly neglected. It is clear that understanding seizure initiation is essential for developing new therapies, but understanding seizure cessation, could also open up entirely new approaches in designing new treatments. In addition, although often overlooked, electrographic patterns found at seizure termination may hold valuable information about the underlying neurophysiology. Indeed, an improved understanding of seizure termination may be harnessed to allow for earlier seizure termination through novel anti-seizure medications or neuromodulatory techniques.^4,5^ Finally, an understanding of how seizures terminate may also provide insight into why some seizures terminate on their own, while others spiral into *status epilepticus*.

While the literature on seizure termination is sparse, there appear to be three theories about how seizures end. The first of these focuses on how metabolic processes change gradually during a seizure, resulting in the termination of the seizure. Following a seizure that shows sustained paroxysmal discharges and neuronal activity, a change in metabolic activity is likely and it is not surprising that metabolic mechanisms may lead to seizure termination.^6^ A second theory on the mechanisms underlying seizure termination involves active increases in inhibition coming from some structure(s) working as a pacemaker^7^. Alternatively, a third theory suggests that seizure termination is associated with a transient increase in network synchrony^8–12^ and that termination is an emergent property of that network level change. None of the proposed mechanisms, however, have been proven for human epilepsy, and more importantly to date no study has considered seizure termination in the context of different seizure types and different patterns of seizure spread.

At first glance, we might assume that all seizures terminate in the same way, especially when compared with seizure initiation wherein ictal activity gradually progresses and often spreads to the entire brain. However, this assumption is an oversimplification and is ultimately inaccurate. While some seizures do terminate simultaneously across the brain (synchronous termination), others do not^13,14^. Seizures in this latter category have been studied only as the subject of seizure onset lateralization (e.g., Trinka *et al*.^13^). In this study, differences in termination patterns were only considered between the two hemispheres, even though seizures may terminate at different times within the same hemisphere.

Seizures can also be differentiated by their bursting patterns. Some seizures exhibit a termination pattern that resembles those found in recordings of neural dynamics in anesthetized patients or those with impaired consciousness.^15,16^ This pattern, being sometimes referred to as “interclonic interval” or “clonic bursts”^16^, is characterized by bursts of high amplitude, high frequency activity or spiking that are interrupted by a period of suppressed activity with very low amplitude. That said, this pattern has received limited coverage in the literature, and it is unclear whether such burst suppression activity can be found in all seizures. We propose that these differences in bursting and termination pattern can be explained by the individual neuronal mechanisms that give rise to two broad classes of seizure: (a) those whose activity is confined to a local area in the brain (focal), and (b) seizures that are secondarily generalized, whose activity propagates to widespread areas and may at times recruit most regions in both hemispheres.

The primary goal of this study, therefore, was to quantify the relationship between patterns at seizure termination and seizure focality, with an eye towards establishing a baseline for mechanistic studies. More specifically, we sought to evaluate the incidence of different burst patterns at seizure termination in four different seizure categories, distinguished by their termination timing (synchronous versus asynchronous) and spread of neuronal activity (focal versus secondarily generalized). We hypothesize that certain bursting patterns (burst suppression versus continuous bursting) manifesting at the termination of both focal and secondarily generalized seizures not only reflect the involvement of different neural circuitry, but also can be indicative of the spread of network recruitment that eventually leads to seizure termination.

### 2 Materials and methods

### 2.1 Data Acquisition and seizure classification

We analyzed seizures recorded from patients with medication-refractory epilepsy who underwent clinical monitoring to locate their seizure onset zone at Massachusetts General Hospital from 2006 to 2019 and at Brigham and Women’s Hospital from 2009 to 2018. Patients were implanted with depth electrodes and/or grids and strips, and their electrode placement was determined by the clinical team independent of this study. All data acquisition and analyses in this study were approved by the Institutional Review Board (IRB) covering the two hospitals (Mass General Brigham and Partners Human Research Committee).

Data were recorded using XLTEK clinical EEG equipment (Natus Medical Inc., Oakville, Canada), Natus Quantum (Natus Medical Incorporated, Pleasanton, CA), or a Blackrock Cerebus system (Blackrock Microsystems). The data were sampled at a range of frequencies from 250 Hz to 2 kHz and were subsequently downsampled to 200 Hz for further analysis.

Seizures were converted into a format compatible for analysis in MATLAB (R2020a; MathWorks) and the data were reviewed (bipolar montage) using the FieldTrip browser.^17^ Epileptologists, blind to this study, identified the seizure onset regions based on EEG reading criteria. We excluded from analysis all seizures from a patient if either: a) the patient’s seizure onset area could not be identified; b) more than three seizure onset areas were identified; or c) none of their seizures lasted more than 30 s. We reviewed and visually classified 710 seizures from 104 patients. Each seizure was labeled according to their focality, termination pattern, and burst pattern. First, seizures were labeled as focal if the ictal activity could only be seen in one recorded area and did not propagate to other brain regions or the contralateral side; otherwise, the seizure was classified as secondarily generalized, meaning that the seizure started in one or multiple area(s) and later propagated to other areas in the brain (Fig. 1-3). Next, we labeled seizures based on their termination patterns: synchronous termination (ST), where seizure activity ended in all channels simultaneously within one second, or asynchronous termination (AT), where seizure activity terminated on some channels while continuing on others. To account for seizure count variability across patients, we randomly selected one representative seizure for each unique (focality, termination pattern, onset zone) label combination from the seizure set for each patient, after excluding those deemed to have excessive noise or persistent, high-amplitude artifacts. Seizures that developed into *status epilepticus* (*N*=7) were excluded from further analysis. The seizure end-time and patterns were identified by two reviewers and discussed until agreement was reached. Where agreement could not be reached, the seizure was marked as unclassifiable (*N*=14) and excluded from further analysis. This resulted in a total of 207 seizures across all patients. Finally, seizures received an additional label based on their burst pattern at termination: a) seizures with synchronous (phase-locked) burst suppression activity across more than 75% of channels at seizure termination (s-BS). b) seizures that show burst suppression activity before seizure termination in most channels, but the burst suppression activity was synchronous in less than 75% of the channels (a-BS), c) seizures with continuous bursting patterns at termination in all channels with ictal activity (CB). The burst suppression pattern has been referred to as “interclonic interval” or “clonic bursts” in other studies.^16^ Here, to correspond with the terminology used in encephalography research and to simultaneously acknowledge the similarity between this pattern and the “burst suppression” pattern, we use the term “burst suppression”.

**Figure 1.**
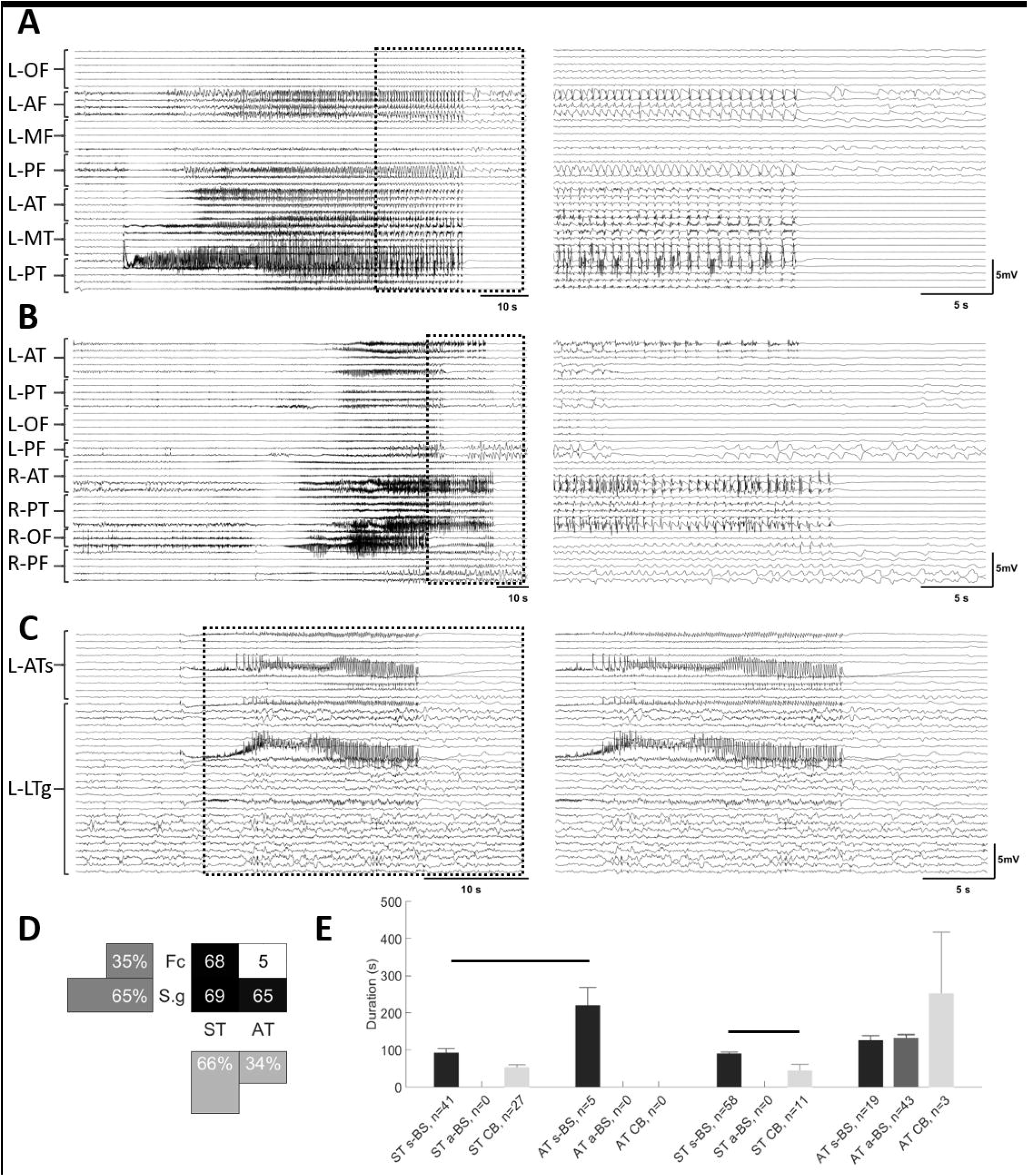
Seizures exhibit different patterns at termination. Examples of seizures with different focality and termination patterns. Seizure traces from 10 s before the onset until 10 s after their termination (A-C, left panels). An expanded view of these traces (A-C, right panels) shows the last 20 s of each seizure and the 10 s post termination period. This 30 s period is identified on the left with dashed boxes. **A)** Seizure with onset in left posterior temporal area with synchronous termination (ST). In the left panel, note the burst suppression activity before the termination. **B)** Seizure with onset in right anterior temporal area that propagates to the rest of the b rain, terminating in some channels while continuing in others (AT). **C)** Seizure that starts focally in left anterior temporal area and remains focal for the duration of the seizure with synchronous termination (ST) with bursting activity at termination. Seizures in each group were further classified into three subgroups: seizures with synchronous burst and suppression activity across almost all channels at the end of the seizure (s-BS, A), and seizures that show burst and suppression activity before seizure termination in most channels, but with burst suppression activity not synchronized across all channels (a-BS, B), seizures with continuous bursting patterns (CB, C). **D)** A distribution of seizures based on focality and the termination pattern (ST and AT). **E)** The duration of seizures in different groups. In both focal and secondarily generalized groups, CB seizure had significantly shorter duration compared to s-BS seizures. Horizontal bars indicate significance between the pairs (p<0.05). L: left; R:right; OF: orbitofrontal; AF: anterior frontal; MF: middle frontal; PF: posterior frontal; AT: anterior temporal; MT: middle temporal; PT: posterior temporal; ATs: anterior temporal strip; LTg: lateral temporal grid; Fc: focal; S.g: secondarily generalized.

### 2.2 Average of absolute energy and burst suppression detection

Seizures were visually analyzed to identify the channels involved in their propagation. The channels that exhibited ictal activities were selected to identify their changes in the average absolute energy (AAE) and burst suppression ratio (47.6±3.24 channels per seizure; min=2; max=201). All seizures were analyzed from 10 s before the seizure-onset until 10 s after the seizure had terminated.

To calculate the AAE of each seizure, each bipolar channel with seizure propagation was band-pass filtered from 2 Hz (to remove the effect of DC changes) to 80 Hz. The absolute value of the amplitude of each channel was measured and averaged over all channels with propagation to represent the changes in the energy of each seizure (Fig. 4A).

**Figure 4.**
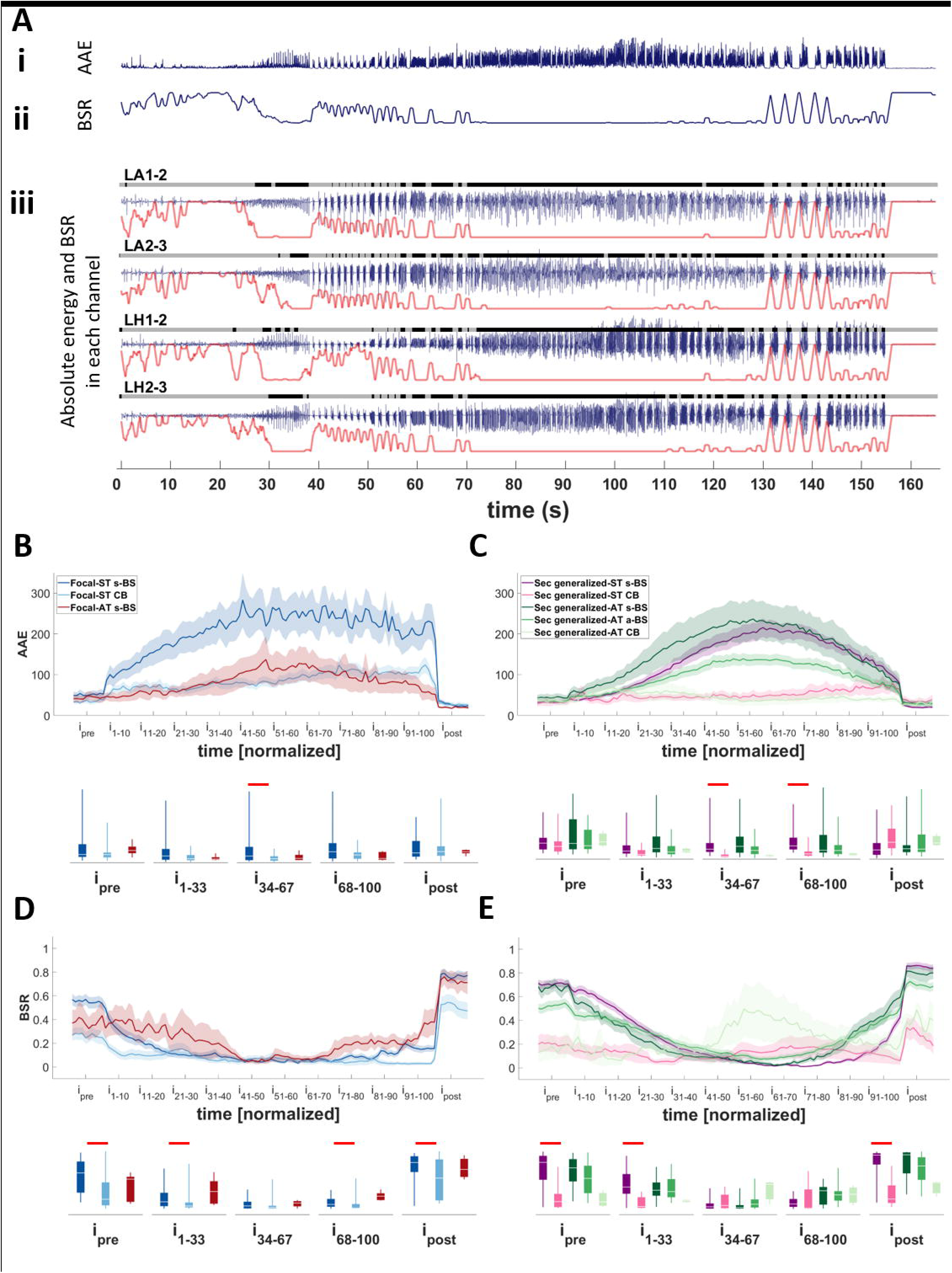
Evaluating AAE and BSR in different seizure types. **A)** An example of a focal seizure that starts in mesial temporal areas and remains focal throughout the seizure duration. Ai) The average absolute energy (AAE) of seizure calculated by averaging the absolute amplitude of channels with seizure spread (four depicted channels for this seizure). Aii) The average value of burst suppression ratio (BSR) of all channels with seizure sprea d. Aiii) For each channel burst (black) and suppression (gray) intervals were identified over time. Note the gray and black bars on top of the signal trace (blue trace) of each channel. The value of the BSR was then calculated for each channel over time (red trace underneath each signal of each channel). These values were then averaged for all channels with seizure activity (here only four channels exhibited seizure activity) **B and C)** Changes in the AAE over time in focal (B) and secondarily generalized (C) seizures with synchronous and asynchronous termination. Note that in both groups the seizures with CB activity at the termination maintain lower energy throughout the seizure compared to the seizures with BS activity. **D and E)** Changes in the BSR in in focal (D) and secondarily generalized (E) seizures with synchronous and asynchronous termination. The box-and-whisker plots underneath the traces indicate summarized measures (AAE or BSR) within different time periods. Traces in B-D indicate averages with a spread of plus/minus one standard error (shaded area). Horizontal bars indicate significance between the pairs (p<0.05). LA: left amygdala; LH: left hippocampus; ST: synchronous termination; AT: asynchronous termination; s-BS: synchronous burst suppression; a-BS: asynchronous burst suppression; CB: continuous bursting; i: interval.

The burst suppression ratio (BSR) was computed using an automated method adapted from Westover *et al*.^18^ Prior to evaluation, the band-pass filtered signals were normalized to account for amplitude differences. Normalization was performed by subtracting the mean of the signal from the signal and dividing by its standard deviation. The automated method then labels each sample as either burst or suppression. Briefly, the method uses the previous data in each signal and applies the following equations:

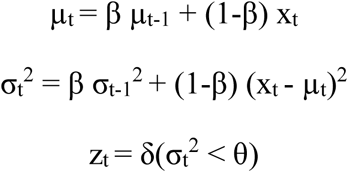

Here, x_t_ is the value of the normalized signal of one channel at time t, µ_t_ and σ_t_^2^ are current values of the recursively estimated local mean and variance, respectively. Finally, z_t_ is an indicator function that labels each data point as either a burst (zero) or suppression (one). The value of β determines the balance between the effect of recent and past data; this was set to the value obtained from previously trained data (β = 0.9534)^18^. The classification threshold θ (i.e., the value above which a data point should be classified as burst) chosen by Westover *et al*.^18^ was evaluated using data recorded from patients in the Intensive Care Unit (ICU) whose brain activity was relatively ‘quiet’ (i.e., with minimal fluctuations in EEG/voltage). We determined through a preliminary analysis that this value of θ is not sufficient to classify the data recorded from epileptic data. We therefore evaluated our dataset visually with values of θ = 0.1, 0.5, and 1.75. The choice of theta was made by two experts who reviewed selected intervals to identify burst and suppression using each possible theta value. The value of θ = 0.1 was selected to reliably identify ictal burst and suppression.

The BSR for each channel was evaluated as the proportion of suppression-labeled samples in a moving window (1 s duration, no overlap). The BSR for each seizure was evaluated as the average over all channels that exhibited the ictal activity (Fig. 4A).

### 2.3 Localizing the region for each contact with seizure propagation

We identified the anatomical location of all bipolar contacts (see: Salami *et al*.^3^ for more detail) that exhibited ictal activity and were determined to be involved with seizure propagation. The location was identified using a combined volumetric and surface registration.^19,20^ Channels from different regions were classified into five distinct groups based on proximity and structural similarity: Region 1) hippocampus; Region 2) mid-temporal structures; Region 3) lateral-temporal area; Region 4) frontoparietal including pre-central, post-central and inferior parietal; Region 5) all other areas.

### 2.4 Functional connectivity and network dynamics

Functional connectivity was measured utilizing linear cross-correlation strength between channels. Connectivity was evaluated on low-pass filtered data (200 Hz cutoff) using a moving window (1 s duration, 0.5 s overlap) within an interval ranging from 10 s before the start of the seizure (pre-seizure period) until 10 s after seizure termination (post-seizure period). The functional connectivity was measured between all intracranial recording channels that contained no artifacts for each seizure (98.8±3.57 channels per seizure; min=26; max=205) and was not limited only to the channels of seizure spread. Using cross-correlation to measure linear coupling between bipolar channel pairs, each bipolar channel was considered as a node and connectivity was measured between each pair. We used a bootstrap method to assess the statistical significance of each connectivity measurement^21^, such that an edge would only be assigned to each pair of nodes with sufficiently strong coupling, irrespective of direction. This resulted in the creation of a connectivity network (indicating the presence or absence of an edge between nodes) for each moving window, which was analyzed using network density (Fig. 7B). This measure indicates the strength of node connectivity in the network and is calculated as density=NE/NPE, the number of edges (NE) as a fraction of the total number of possible edges, NPE = n(n-1)/2, where n is the number of nodes, or channels.

**Figure 7.**
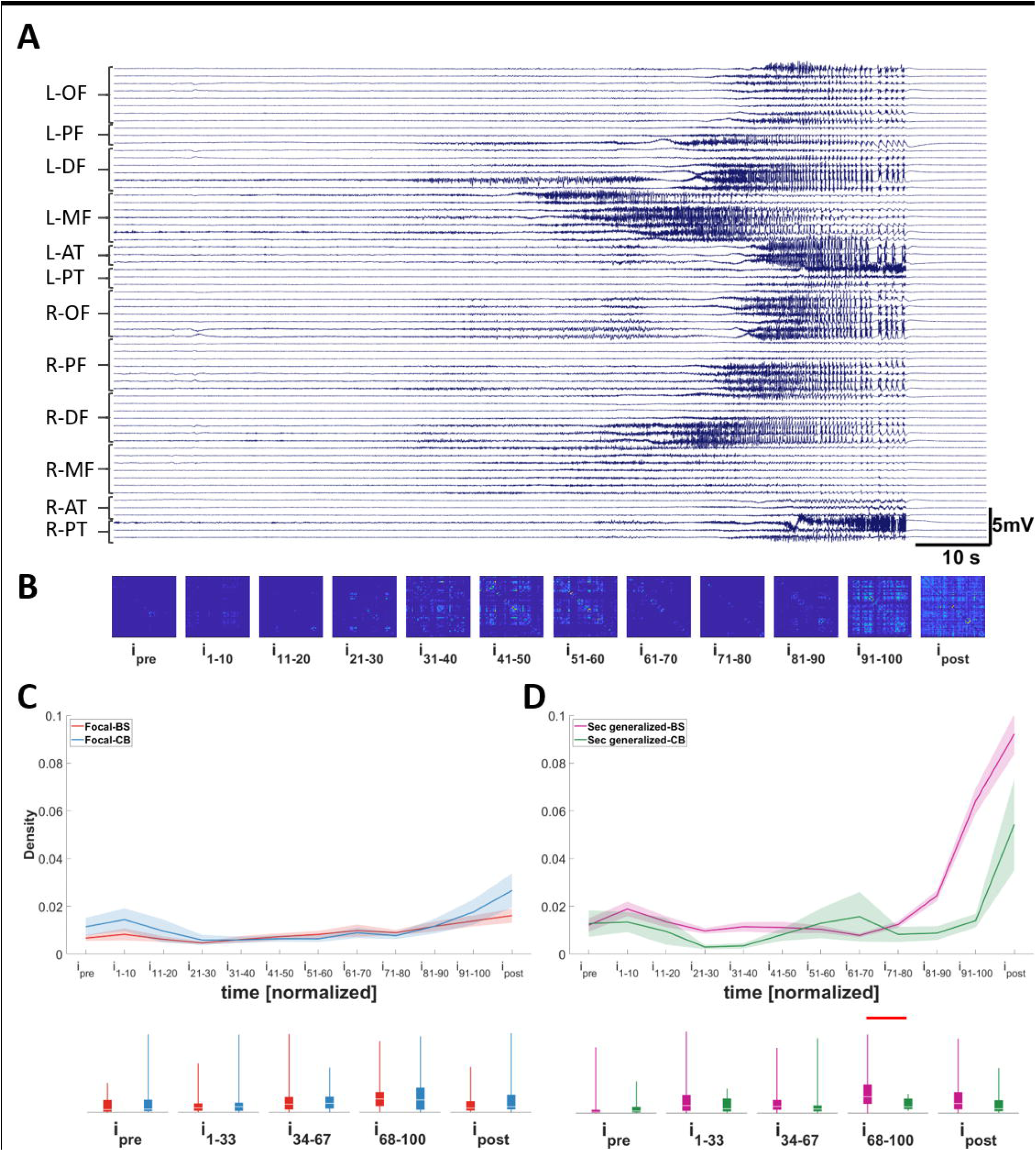
Changes in network density in different seizure types. **A)** Seizure with onset in left posterior temporal area; synchronous termination (ST). **B)** Average connectivity matrix over time for the seizure shown in A. **C and D)** Differences in average density between seizures with different bursting activity at their termination in focal (C) and secondarily generalized (D) seizures. Note that there is a transient increase in the density over time in all types of seizures. The density increases significantly in secondar ily generalized seizures with burst suppression activity at their termination indicating an increase in the synchrony in these seizures. Traces in C-D indicate averages with a spread of plus/minus one standard error (shaded area). The box-and-whisker plots underneath the traces indicate summarized measure (density) within different time periods. Horizontal bars indicate significance between the pairs (p<0.05). L: left; R:right; OF: orbitofrontal; PF: posterior frontal; DF: dorsal frontal; MF: middle frontal; AT: anterior temporal; PT: posterior temporal; BS: burst suppression; CB: continuous bursting; i:interval.

### 2.5 Statistical analysis

We used the Kruskal-Wallis test to identify significant differences between the duration of seizures of different groups. This was followed by the Tukey-Kramer method for multiple comparisons with the level of significance set at *p*=0.05. To compare measures of AAE, BSR and network density between seizures, the differences in seizure duration need to be taken into account. The duration of each seizure was divided into 100 subintervals of equal duration so as to normalize the time during the seizure. The value representing each interval was the average of the measures of all samples within each interval. All seizure measures were then averaged and plotted in this normalized time scale. In addition, we considered a 10 s period before the seizure onset (pre-seizure period) and a 10 s period after the seizure ending (post-seizure period). To test differences between the groups over time, seizures were compared in five time periods: pre-seizure, beginning of the seizure (subintervals 1-33), middle of the seizure (subintervals 34-66), end of the seizure (subintervals 67-100), and post-seizure periods. For each time period, the Kruskal-Wallis test was used to identify significant differences in AAE and BSR between seizures with similar focality but different termination patterns. The Tukey-Kramer method was used for multiple comparisons with the level of significance set at *p*=0.05. To compare the differences between network density of seizures with different bursting patterns, we used Wilcoxon rank-sum test with the level of significance set at *p*= 0.05. Results in the text and bar graphs are reported as mean ± standard error of the dataset across patients or seizures.

### 2.6 Data availability

The data of this study are available upon request from the corresponding author. The data are not publicly available due to their containing information that could compromise the privacy of the participants.

## 3 Results

### 3.1 Individual patients can have seizures with different termination types

We analyzed a total of 710 seizures recorded from 104 patients. In our entire dataset (*n*=710), seizures with asynchronous termination (AT) appear frequently (18%). In the seizures selected for analysis, most (65%) were secondarily generalized. Of the 35% of seizures that were focal, most had synchronous termination (ST; 93%). After labelling the seizures from each patient based on their focality and termination characteristics, representative seizures for that patient were selected randomly for every unique combination of focality, termination pattern, and onset location for that patient (see Methods). This resulted in a total of 207 seizures for analysis across all patients (2±0.1 seizures per patient; min=1; max=7). In our analyzed seizure pool, 51% that secondarily generalized had ST, with the rest having AT (Fig. 1D).

Even in seizures with AT, the ictal activity typically appeared to end abruptly within different regions. Unlike the gradual recruitment of regions at seizure initiation, where a seizure spreads slowly and continuously across the brain, during seizure termination, the seizure either ends across all channels at the same time (ST) or the termination happens in groups of channels belonging to the same region (AT; Fig. 2).

**Figure 2.**
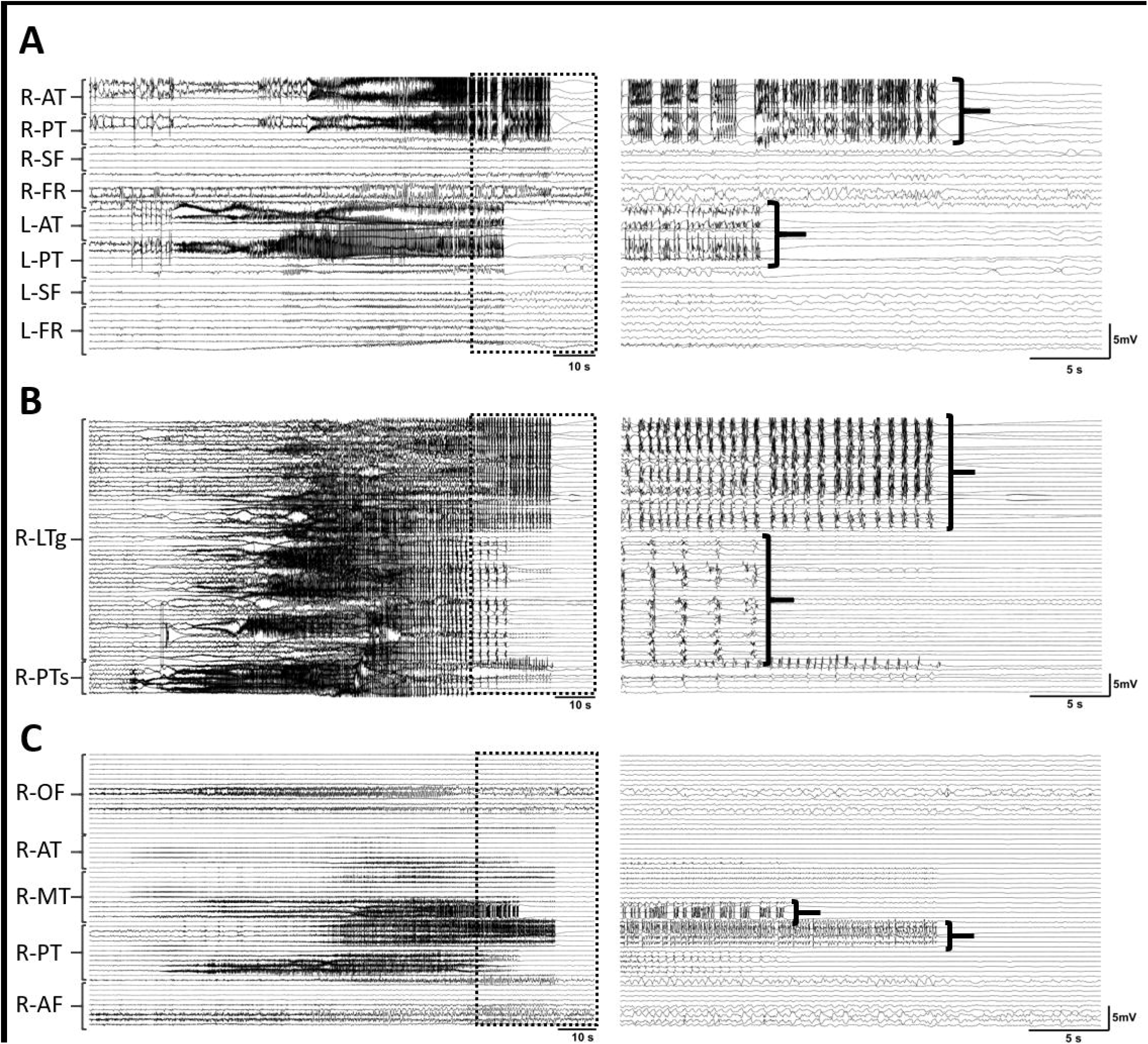
Ictal activity in asynchronous termination, unlike seizure initiation and spread, appears to end abruptly within different regions. Three seizures with asynchronous termination and asynchronous bursting patterns (a-BS) from three different patients with onset in left posterior temporal **(A)**, right lateral temporal **(B)**, and right lateral temporal **(C)** regions. Seizures are presented as per Fig. 1. Note that ictal events from different regions end in groups. L: left; R:right; AT: anterior temporal; MT: middle temporal; PT: posterior temporal; SF: sub-frontal; FR: frontal; OF: orbitofrontal; AF: anterior frontal; PTs: posterior temporal strip; LTg: lateral temporal grid.

Among the 104 patients, six had *status epilepticus* only or seizures whose termination could not be identified (unclassifiable). Fifty-nine patients had only one type of termination (81% ST-only and 19% AT-only), five of which had at least one extra unclassifiable seizure or seizures that lead to *status epilepticus*. Thirty-nine patients had both types of ST and AT, and four had at least one unclassifiable seizure or a seizure that lead to *status epilepticus*.

### 3.2 Seizures manifest different bursting patterns at termination

In addition to focality and termination pattern, seizures were also distinguished by their bursting patterns. We identified three subgroups: seizures with synchronous burst and suppression activity across almost all channels at the end of the seizure (synchronous burst suppression or s-BS; Fig. 1A), seizures that show burst and suppression activity before termination in most channels but with burst suppression activity not synchronized across all channels (asynchronous burst suppression or a-BS; Fig. 1B), and seizures with continuous bursting with no intervals of suppression (continuous bursting or CB; Fig. 1C). All focal seizures with ST (*N*=68) either had CB (40%) or s-BS (60%) burst patterns. This was also true of secondarily generalized seizures with ST (*N*=69; 16% CB, 84% s-BS). Only five seizures manifested as focal with AT, with all of these having the s-BS pattern. Only secondarily generalized seizures with AT manifested with all three burst patterns (66% a-BS, 30% s-BS, 4% CB). Focal seizures with ST and s-BS patterns had significantly shorter durations than those of AT (Fig. 1E). Secondary generalized seizures with s - BS patterns were significantly longer than seizures with CB patterns in the ST group (Fig. 1E).

It is noteworthy that in some seizures with a-BS and s-BS ending patterns, the mesial temporal area occasionally exhibited bursting activity while other regions were suppressed. This type of activity was seen mostly in the hippocampus, but other times appeared in amygdala and adjacent regions (Fig. 3).

**Figure 3.**
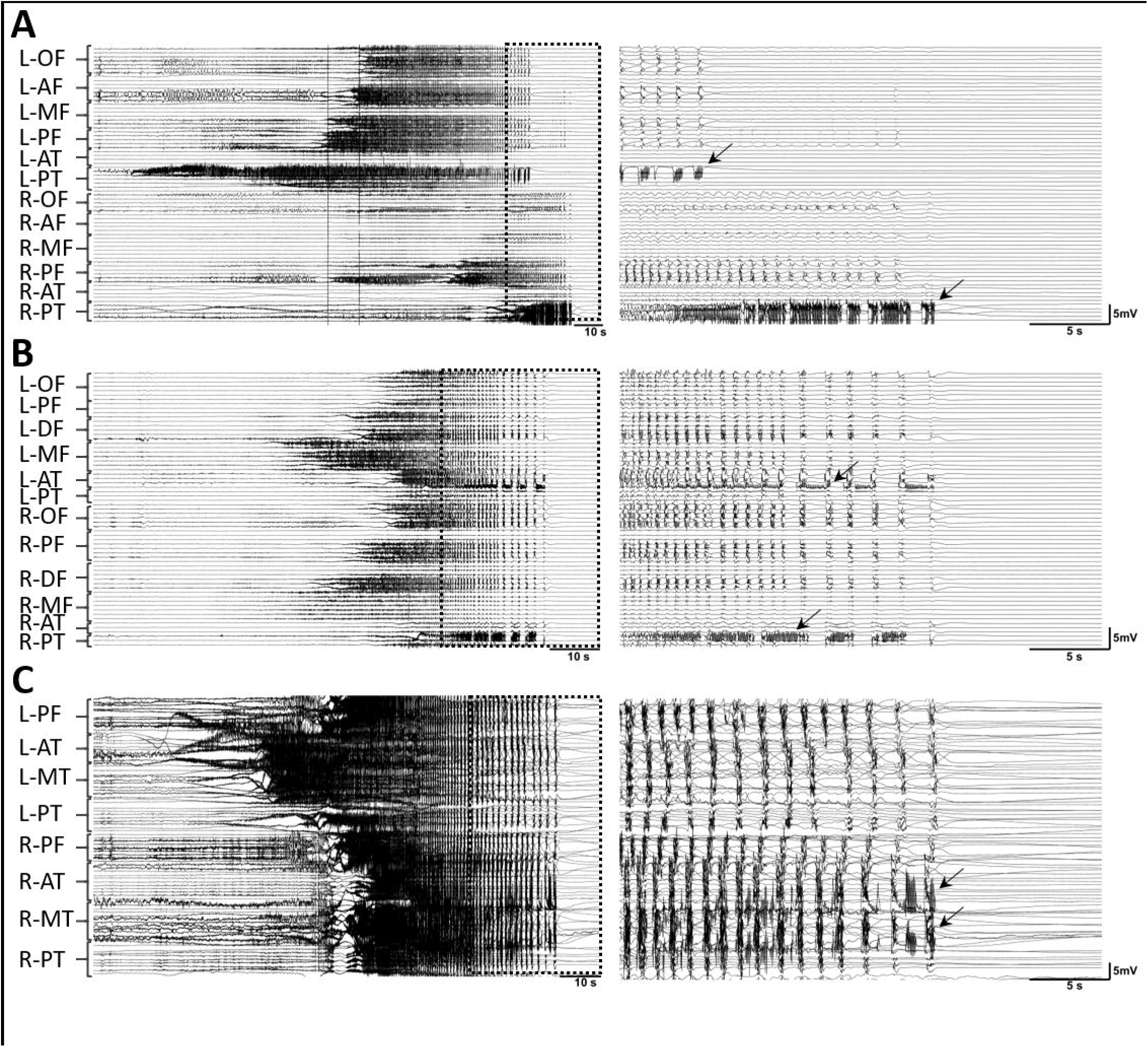
In some seizures with burst suppression activity at their termination, the mesial temporal area exhibits bursting activity while other regions are suppressed. Three seizures with burst suppression activity from three different patients. Seizures are presented as per Fig. 1. **A)** Seizure with an onset in left posterior temporal and asynchronous termination. The arrows are indicating the bursting activities in the left and right hippocampus while the rest of the regions are suppressed. **B)** Seizure with an onset in left middle frontal that propagates to the rest of the brain. The arrows are showing the bursting activity in left and right hippocampus. **C)** Seizure that starts in left posterior temporal area before spreading to the rest of the brain. The arrows showing the bursting activity in the right hippocampal regions. L: left; R:right; OF: orbitofrontal; AF: anterior frontal; MF: middle frontal; PF: posterior frontal; AT: anterior temporal; M T: middle temporal; PT: posterior temporal; DF: dorsal frontal.

### 3.3 Seizures with burst suppression patterns at termination show higher amplitudes

Signal amplitude may serve as a measure of synchrony in the network.^22^ To compare the differences in synchrony with this measure we calculated the changes in average absolute energy (AAE) during the seizure. For each seizure, we first computed the absolute energy over time for each channel, then averaging over all channels that showed ictal activity (Fig. 4A, top trace) to obtain the AAE. We then summarized each energy average as a 100-element sequence in which the first and last values represent the AAE at the beginning and end of the seizure, respectively (see Methods). This normalization over time allowed us to directly compare different seizures which naturally have different durations (Fig. 1E). The average energy traces of focal seizures with synchronous and asynchronous endings are shown in Fig. 4B, and those of secondarily generalized seizures with both synchronous and asynchronous endings are shown in Fig. 4C.

There were no differences in absolute energy between seizures with synchronous and asynchronous endings. However, in both groups, the seizures with s-BS reached significantly higher amplitudes during the seizure compared to seizures with CB in both focal (Fig. 4B; *p*<0.03) and secondarily generalized groups (Fig. 4C; *p*<0.0005).

### 3.4 Seizures which terminate with burst suppression exhibit more suppression during pre- and post-ictal periods

The burst suppression ratio (BSR) indicates the proportion of time in which a signal is suppressed and is equal to zero (one) if the entire period consists of bursting (suppression). The BSR for each seizure was computed in a similar fashion to AAE: the BSR for each channel containing ictal activity was averaged (Fig. 4A) and normalized in time as a 100-element sequence.

We compared seizures with ST and AT in both focal (Fig. 4D) and secondarily generalized seizures (Fig. 4E). We did not identify any significant differences in BSR changes between seizures with synchronous and asynchronous termination in either group (*p*>0.05). In all groups the pre-seizure period featured bursting and suppressed activity, with bursting activity increasing (and BSR falling) on seizure onset. Towards the end of the seizure, the bursting activity became interrupted by periods of suppression and the BSR value started to increase until termination. In seizures that did not feature suppressed activity before termination (seizures with CB patterns), the BSR remained relatively stable from pre-seizure to post-seizure. Interestingly, the BSR in seizures with CB patterns tended to be significantly lower than that of seizures in the same group (in terms of focality and synchronous or asynchronous termination) but with s-BS activity in the pre- and post-seizure periods in both focal (Fig. 4D; *p*<0.03) and secondarily generalized seizures (Fig. 4E; *p*<0.00003). It is important to note in the group of secondarily generalized seizures with AT there were only three seizures exhibiting the CB pattern in our dataset. Therefore, even though the BSRs of these seizures were different from those with the BS pattern, this difference did not reach significance (Fig. 4E).

### 3.5 Hippocampus has high amplitude in secondarily generalized seizures with burst suppression pattern

In each seizure, the channels that exhibited seizure spread were located in different regions, which were classified into five distinct groups based on proximity and structural similarity: Region 1) hippocampus; Region 2) mid-temporal structures; Region 3) lateral-temporal area; Region 4) frontoparietal including pre-central, post-central and inferior parietal; Region 5) all other areas. For each seizure type, we compared the AAE and BSR changes over time in different regions to determine whether all regions behave in similar ways over the duration of the seizure. To perform this comparison, channels situated in the same region were grouped and averaged together for each seizure. We compared only the activity in the four major regions where seizures occurred most frequently. Some areas of interest (such as thalamus) were rarely recorded from in our selected patients. Thus, all areas that were classified into the ‘leftover’ Region 5 group were excluded from this analysis due to low case count. In focal seizures with and without burst suppression pattern at termination, hippocampal regions demonstrated higher AAE towards the end of the seizure; however, this difference did not reach significance (Fig. 5A and 5B; *p*=0.055 for BS seizures and *p*=0.82 for CB seizures). In seizures with secondarily spread that exhibited burst suppression pattern at their termination, hippocampal regions showed significantly higher AAE during the seizure (Fig. 5C; *p*<0.05). There were no significant differences in AAE between any of the regions in secondarily generalized seizures with CB patterns (Fig. 5D; *p*>0.05).

**Figure 5.**
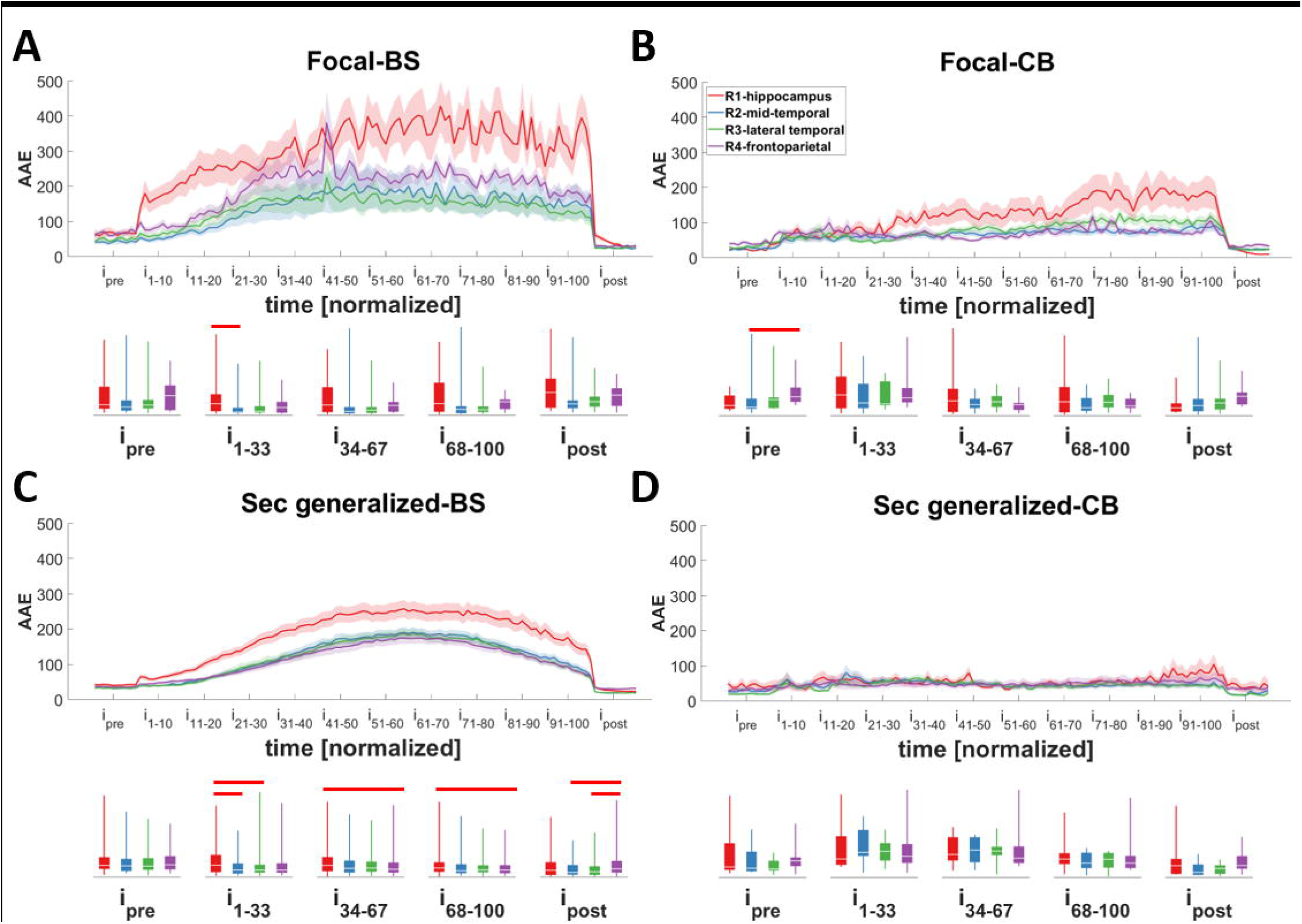
Changes in the average absolute energy (AAE) of different regions during different seizure types. Seizures were classified into different groups based on their focality: **A and B)** focal; **C and D)** secondarily generalized, and the bursting pattern at their termination: **A and C)** asynchronous burst suppression (a-BS) and synchronous burst suppression (s-BS); **B and D)** continuous bursting (CB). Hippocampus tend to have higher AAE in seizures with burst suppression activity at their termination (A and C). This difference has reached significance in multiple intervals in secondarily generalized seizures with burst suppression (a-BS and s-BS). Traces indicate averages with a spread of plus/minus one standard error (shaded area). The box- and-whisker plots underneath the traces indicate summarized measure (AAE) within different time periods. Horizontal bars indicate significance between the pairs (p<0.05; i: interval).

When comparing BSR over time in seizures with different focality and termination patterns, we found that most regions behave similarly except for frontoparietal regions. Before the seizure starts, all regions had a baseline consisting of both burst and suppression activities; however, frontoparietal regions exhibited more bursting activity (and less suppression), especially compared to hippocampus and mid-temporal areas during pre-seizure periods and this difference reached significance in some seizure types. This behavior persisted after the seizure end where all regions entered a suppressed period, while frontoparietal regions exhibited more bursting activity (Fig. 6B-D; *p*<0.05).

**Figure 6.**
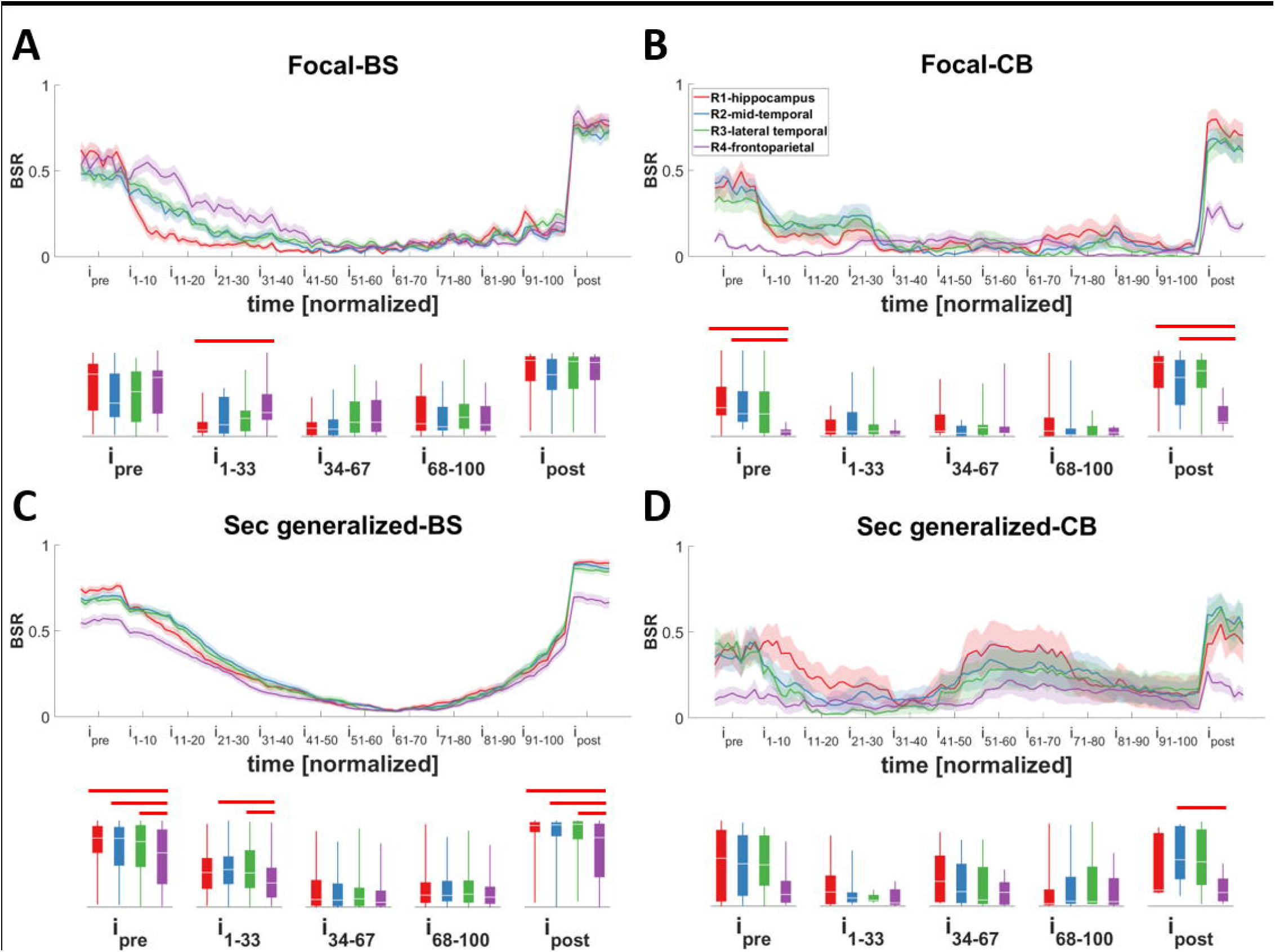
Changes in the burst suppression ratio (BSR) of different regions during different seizure types. Seizures were classified into different groups based on thei r focality: **A and B)** focal; **C and D)** secondarily generalized, and the bursting pattern at their termination: **A and C)** asynchronous burst suppression (a-BS) and synchronous burst suppression (s-BS); **B and D)** continuous bursting (CB). Note that most regions behave similarly except for frontoparietal regions. Except for focal-BS seizures, before the seizure starts and after the seizure ends, frontoparietal regions show more bursting activity (and less suppression) especially compared to hippocampus (B, C, and D). Traces indicate averages with a spread of plus/minus one standard error (shaded area). The box-and-whisker plots underneath the traces indicate summarized measure (BSR) within different time periods. Horizontal bars indicate significance between the pairs (p<0.05; i: interval).

### 3.6 Network density at the termination of seizures with burst suppression activity is higher than the seizures with continuous bursting

In both CB seizures and BS seizures, we did not identify any significant differences in AAE or BSR between the seizures with synchronous or asynchronous termination; however, CB seizures tend to have lower AAE and a significantly lower BSR over the entire course of the seizure compared to BS seizures (Fig. 4C-D). To better understand these differences, we evaluated the changes in the network connectivity of CB and BS seizures. In focal and secondarily generalized seizure groups, we combined all the seizures with burst suppression patterns (a-BS and s-BS) together and compared them with CB seizures. For seizures with focal onsets, no differences between the density of functional connectivity network between the two types were identified (Fig. 7C). This result was somewhat predictable; the density of functional connectivity in focal seizures tends to be low since the areas that are involved in seizure propagation are confined to a small region of the brain. However, in secondarily generalized seizures, those that demonstrate a burst suppression pattern at termination (a-BS and s-BS) exhibited an increase in network density with progression of the seizure (Fig. 7D; p=0.002). The peak network density of a-BS and s-BS seizures (occurring in the last interval of the seizure) was significantly higher than that of CB seizures. These findings show that there is more synchronization in seizures with BS patterns compared to seizures with CB patterns.

## 4 Discussion

Seizure termination occurs through complex mechanisms that are influenced by a variety of factors though many of these factors and mechanisms remain unknown. To address a portion of this unknown territory, our main goal in this study was to devise a classification scheme to describe the various ways in which a seizure may terminate. The electrographic patterns that manifest at seizure termination exhibit less variety than those at seizure initiation and can be grouped into distinct categories. We observed two broad types of termination: synchronous termination (ST) and asynchronous termination (AT). Within each group, we found that termination patterns could be distinguished by their bursting patterns: a) synchronous burst suppression activity in almost all channels at seizure termination (s-BS), b) asynchronous burst and suppression activity before seizure termination (a-BS), and c) continuous bursting patterns at termination in all channels with ictal activity (CB). The main findings of our study are that 1) different seizure termination patterns can manifest within a single patient, even in seizures with similar seizure onsets; 2) most seizures terminate with patterns of burst suppression, regardless of their generalization; 3) seizures that secondarily generalize show burst suppression patterns in 90% of cases; 4) seizures with synchronous and asynchronous termination have similar average of absolute energy (AAE) and burst suppression ratios (BSR); however; 5) those seizures that have continuous bursting activity (CB) have lower AAE during the seizure compared to seizures with burst suppression activity (s-BS); 6) AAE in secondarily generalized seizures with BS pattern recorded from hippocampus tends to be higher compared to those in other regions; and finally, 7) the network density increases with seizure progression (reaching a maximum at termination), with significantly lower densities in CB seizures compared to seizures with BS activity (a-BS and s-BS).

### 4.1 Seizure termination patterns

All seizures analyzed in this study had initiated with at least one focus before either gradually spreading to other regions (secondarily generalized seizure) or remaining confined to one region (focal seizure). While seizure initiation and spread tend to progress gradually, seizure termination is usually abrupt (Fig. 1A-C), even in seizures that exhibit AT (Fig. 1B and Fig. 2). While this feature might be obvious (particularly to encephalographers), its importance is often overlooked. Even in the case of AT, localized groups of channels are seen to terminate together, suggesting that ictal activity, either within a single region or shared across functionally-connected regions, ends all at once (Fig. 2). These results suggest there are clear differences in the processes leading to seizure initiation compared to seizure termination.

We have found that roughly a fifth of seizures in both focal and secondarily generalized types exhibit bursting activity throughout the seizure (CB seizures), and that this activity is sustained until termination when it is abruptly suppressed or reverts to baseline. Although we identified a small number (10%) of secondarily generalized seizures of this nature, the majority of such seizures were focal; in cases where these CB seizures spread to other areas, they propagated to relatively fewer regions. Most secondarily generalized seizures (90%) show a BS pattern at termination. This pattern can be compared to the synchronized pattern seen in seizures of idiopathic generalized epilepsy, where all regions exhibit synchronized voltage fluctuations, suggesting the involvement of thalamocortical activity known to be common in idiopathic generalized epilepsy.^23,24^

While some studies have used the DC shift at seizure offset to discriminate between seizures ^25^, we did not do this in our study. Most seizures investigated in this study were recorded with non-DC amplifiers and as such we could not investigate this relationship. However, in our previous study of seizure initiation ^3^, we found that the relationship between DC shifts (or infraslow activity; ISA) and seizure generation is unclear. It is possible that seizures exhibit changes in DC shift at the beginning of the seizure followed by a return to baseline when the seizure ends. Changes in DC shift are believed to reflect changes in extracellular ionic concentration, which are thought to be involved in seizure termination.^26^ It would be interesting to see whether there is a relationship between (say) extracellular potassium ions and the seizure types introduced in this study, as well as whether certain ion concentrations manifest during specific seizure types.

Other studies have proposed differentiating seizures by their temporal dynamics.^25^ Unlike their classification, which focuses on changes in frequency or amplitude in one channel before seizure termination, we propose in our study a different classification method that considers the interaction between different channels. While there are advantages in considering seizure termination through either method, we believe that our classification can help understand how seizure spread to different regions can influence the specific pathways by which they terminate.

### 4.2 Differences in absolute energy and burst suppression activity indicate differences in synchronization

We propose that seizures with continuous bursting activity (CB seizures) have a different ending mechanism compared to seizures with burst and suppression termination patterns (a-BS and s-BS). This is based on differences in the AAE during seizure progression. More specifically, we observed that CB seizures have significantly lower AAE compared to those seizures with synchronous burst suppression activity (s-BS). At the LFP level, the amplitude indicates the level of synchrony between focal groups of neurons that contribute to the generation of the signal.^22^ If more neurons activate simultaneously, the recorded signal will register with a higher amplitude. In most seizures, at the beginning of the seizure a high-frequency, low-amplitude pattern can be identified whose amplitude is relatively small at initiation.^3^ From this point, the amplitude gradually increases as the seizure progresses and more networks of neurons are recruited. Since the AAE in CB seizures remains low, we hypothesize that their recruited networks are relatively small and do not change over the course of a seizure. If this were true, it would suggest that a different mechanism is responsible for the initiation, spread and termination of CB seizures compared to seizures with burst suppression activity (a-BS and s-BS).

### 4.3 Network connectivity and network density

We also found differences in the network density of CB compared to seizures that present intervals of suppression between bursts (a-BS and s-BS). As discussed earlier, we believe that this activity is induced by increased synchronization between different brain regions. We investigated this hypothesis by evaluating changes in network density over time, finding that the network density increases in secondarily generalized a-BS and s-BS seizures as they reach termination. The increase in density (indicating an increase in the number of edges in the network) suggests highly synchronous activity, which supports the idea that synchrony between different areas may influence seizure termination.^10,11^ This finding suggests that global or widespread activity may play a role in seizure termination of seizures with BS activity, and whatever mechanism supports this increased synchrony influences many brain regions at the same time. In a subset of seizures this effect might be delayed in some regions leading to asynchronous termination. This synchronization may not be present in all seizures (e.g., CB seizures) since we believe a certain amount of spread to subcortical regions and to the rest of the brain is necessary for its presence. In fact, we observed a much lower density in focal seizures with both CB and s-BS activity at their termination compared to secondarily generalized seizures.

### 4.4 Variability of patterns at termination implies differences in mechanisms

Our findings suggest that not all seizures terminate in the same way. Different factors can play a role in seizure termination and, depending on the seizure type, the importance of these factors may change. Several metabolic mechanisms are thought to play a crucial role in seizure termination. Among these, changes in pH caused by sustained synaptic transmission ^27,28^, adenosine^29^, and extracellular volume along with neurotransmitter depletion ^30^ as well as changes in ionic concentration^26,31^ have been supported by findings from animal studies. None, however, have been the focus of investigation in human studies, and so it is unclear which of these possible mechanisms are at play. Several studies in both animals and humans have shown that the changes in oxygen or glucose are brief, arguing against their role in termination.^32^ Nevertheless, this explanation cannot account for the development of *status epilepticus* in some seizures. Nor can it, on its own, explain features of seizure termination like synchronization.

We suspect that metabolic factors might work to either speed up or slow down discharges in CB seizures. Alternatively, the synchronous activity exhibited in seizures with BS at the end might be mediated by subcortical regions including (but not limited to) thalamus, which would induce inhibitory effects throughout the brain.^7^ Since most of the seizures with both synchronous and asynchronous termination demonstrate relatively constant bursts of activity (a-BS and s-BS seizures), the mechanisms behind the cessaion of these seizures produces an increase in synchrony between different regions. Thus, metabolic factors alone are unlikely to cause such a synchronous termination across the brain.

Burst suppression activity can be recorded during anesthesia or impaired consciousness.^15^ In the decades since its description^33^, researchers have investigated the neurological correlates of this activity. It seems reasonable to assume that cortical networks enter a brief ‘silent mode’ between bursts, however studies^34,35^ in cats showed that this was not the case for subcortical regions including thalamus and hippocampus. Steriade *et al*. ^34^ showed that at least 30% of thalamic neurons are active during the suppression period. In a later study, Kroeger *et al*. ^35^ demonstrated that hippocampal activity increases with higher levels of network synchrony, a finding that we observed in a subset of our seizures with burst suppression pattern at their termination (Fig. 3).

The thalamus plays an important role during synchronization, as evidenced by various animal studies and human imaging studies.^36,37^ This notion may be further supported by the finding that thalamic stimulation can be used to control seizures.^38^ Since intracranial recording from thalamus is uncommon in epileptic patients, it was not possible to investigate the differences between thalamic and cortical termination activity in our study. However, there are examples of hippocampal activity in our data that show continuous patterns of bursting while the rest of the brain exhibits a suppressed period, during the burst suppression period of the seizure. Further, we showed that hippocampus seizures tend to have higher AAE compared to other regions in secondarily generalized seizures with burst suppression activity at their termination.

As suggested and reviewed by Norden and Blumenfeld^36^, subcortical structures including the thalamus, basal ganglia, and brain stem nuclei may play important roles in synchronizing brain activity between different regions, which may facilitate seizure propagation. Two brainstem nuclei – the dorsal raphe nuclei and locus coeruleus – are known to produce global neuromodulatory effects through the release of their respective neurotransmitters, serotonin and norepinephrine. Serotonin, in particular, is known to have an effect on seizure occurrence, with increases in extracellular serotonin inhibiting both focal and generalized seizures.^39^ In addition, previous studies have demonstrated that the locus coeruleus projects bilaterally ^40^ and its stimulation can attenuate seizures.^41–44^ Further, lesions of the locus coeruleus reduce the possible benefits of vagal nerve stimulation and may even facilitate the pathway to *status epilepticus*.^45,46^

The findings of our study suggest that unlike seizure initiation, there is a limited number of dynamical patterns at seizure termination. This implies that the number of mechanisms at seizure termination are also fairly limited. We anticipate that further study of these termination patterns and their differences may provide useful clues about how to manage different types of seizures, which may vary depending on which regions they reach. Since the networks responsible for synchronization in seizures with burst suppression might vary across regions, different regions should be targeted depending on the specific seizure type. Further work in this area could investigate whether there is a relationship between the electrographic pattern or region of onset^2,3^ and the pattern at termination. If the seizure onset region and pattern can be informative for the recruitment of subcortical regions and their effect on seizure spread and progression, this might get us one step closer towards identifying the regions responsible for synchronous activity, which will be useful in controlling seizures with devices using closed-loop stimulation.^47^

## Acknowledgements

The authors wish to thank Alex Hadjinicolaou, Angelique Paulk, Dan Soper, and Rina Zelmann for their invaluable insight on drafts of this manuscript. We are also grateful to the clinical team, technicians and our participants who selflessly help us further our knowledge of the brain.

## Abbreviations

IRB: Institutional Review Board
ST: synchronous termination
AT: asynchronous termination
s-BS: synchronous burst suppression
a-BS: asynchronous burst suppression
BS: burst suppression
CB: continuous bursting
AAE: average of absolute energy
BSR: burst suppression ratio

## Supplementary Material

None.

